# The development and usability testing of digital knowledge translation tools for parents of children with bronchiolitis

**DOI:** 10.1101/2021.06.21.21259266

**Authors:** Anne Le, Lisa Hartling, Shannon D. Scott

## Abstract

Bronchiolitis is an acute infection of the lower respiratory tract that predominantly affects children less than two years old. Although self-limiting, symptoms of bronchiolitis can be distressing for young children. Research has demonstrated that parents may not have the necessary information to be able to identify bronchiolitis symptoms, resulting in emergency department (ED) visits and hospitalizations. Parents have expressed that they feel unprepared, afraid, and that they lack information on their child’s condition. Digital knowledge translation (KT) tools have the potential to convey complex health information to parents to support their healthcare decision-making needs.

We worked with parents of children with bronchiolitis to develop and evaluate three digital tools on bronchiolitis (whiteboard animation video, infographic, and e-Book). Following prototype completion, usability testing was conducted using iPads in two Alberta ED waiting rooms. Parents were randomized to one out of the three tools. Overall, the tools were highly rated, suggesting that arts-based digital tools are useful in delivering complex health information to parents.

## Introduction

Bronchiolitis, an acute lower respiratory tract infection, often affects children less than two years old and is the most common cause of hospitalization for infants under the age of 1 [1-2]. Often caused by the respiratory syncytial virus (RSV), characteristics of bronchiolitis include acute inflammation, edema, necrosis of epithelial cell linings, and increased mucus production [2-3]. Although bronchiolitis is often self-limiting, small amounts of inflammation may cause respiratory distress due to the small anatomy of children [4]. As a result of the wide variation in disease states, methods of bronchiolitis diagnosis and treatment vary and overuse of unnecessary tests and treatments are frequent [2,5]. Previous research has identified that parents may not always be aware of bronchiolitis symptoms and when to seek care, and as a result, have experienced feelings of fear, isolation, and helplessness [6]. This demonstrates the need for parents to understand the condition and more effective knowledge sources [6].

Research evidence suggests that targeting health consumers such as parents may aid in the development of tools that facilitate understanding and decision making [7-8]. Furthermore, targeting knowledge translation (KT) resources at parents may reduce health care utilization and improve health outcomes [9]. Past literature has demonstrated the power of innovative methods over traditional written information, highlighting the potential for art and narrative-based approaches, such as infographics, when educating patients and families [10-12]. To date, there have been limited tools developed using art forms to educate about bronchiolitis. Existing tools often discuss general respiratory tract issues or care and treatments [13-14]. In addition, these tools commonly target health care providers and specific minority populations [15-16].

As co-directors of Translating Emergency Knowledge for Kids (TREKK), our research programs develop arts-based KT tools that deliver consumer-friendly, evidence-based information for parents. Combining parental narratives and best practice guidelines, we have developed an innovative method to equip parents with information and strategies to manage common childhood conditions (e.g., gastroenteritis, chronic pain, croup, asthma). The purpose of this study was to work with parents to develop and assess the usability of a whiteboard animation video, e-Book, and interactive infographic for bronchiolitis in children.

## Methods

We employed a multi-method study involving patient engagement to develop, refine, and evaluate a whiteboard animation video, e-Book, and interactive infographic for pediatric bronchiolitis (**Figure 1**). Research ethics approval was obtained from the University of Alberta Health Research Ethics Board (Edmonton, AB) [Pro00062904]. Operational approvals were obtained from individual emergency departments and urgent care centres to conduct usability testing.

**Figure 1.**
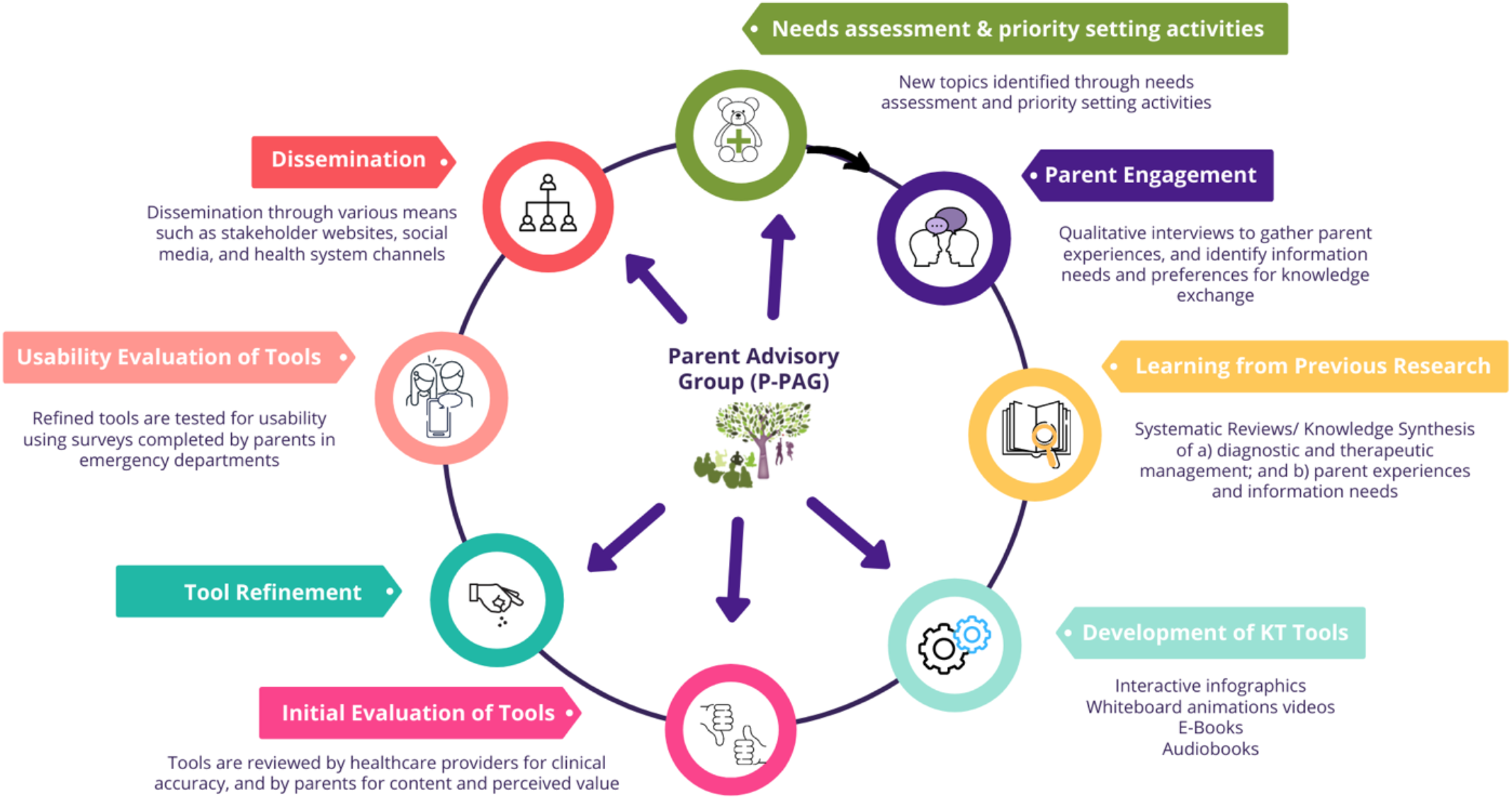
KT Tool Development Cycle.

### Compilation of Parents’ Narratives

Parental narratives were informed through semi-structured qualitative interviews (**Appendix A**) and a systematic review [6, 17-18]. Parents of children who presented to the Stollery Children’s Hospital (Edmonton, Canada) with bronchiolitis were invited to participate. Interviews were conducted by a research coordinator trained in qualitative methodology. Parents (n = 15) were asked to discuss their experiences having a child with bronchiolitis. Concurrently, a systematic review was conducted to synthesize current evidence about experiences and information needs of parents and caregivers managing bronchiolitis. Key findings from the systematic review and qualitative study were used to develop content outlines for a whiteboard animation video, e-Book, and interactive infographic. Results from the systematic review and qualitative interviews are published elsewhere [6, 17-18].

### Intervention Development

Storyboards, scripts, and skeletons were written by the research team and shared with a creative team to develop the e-tool prototypes. The creative team, which included graphic designers, artists, video developers, and authors were selected through a competitive process. Each tool included best available evidence on pediatric bronchiolitis from the TRanslating Emergency Knowledge for Kids (TREKK) Bottom Line Recommendations (BLR) [19]. Information pertinent to management of pediatric bronchiolitis was embedded within the storyline of the video and e-Book, which depicted parents struggling to manage their child’s condition (**Appendices B and C**). Likewise, information on symptoms, treatment options, and when to seek emergency care were included in the infographic (**Appendix D**).

### Revisions

Iterative processes were used to develop the tools. We sought feedback from a variety of different stakeholder groups, including parents, health care professionals (HCPs), and researchers. HCPs were asked to comment on the accuracy of information and evidence, length of the tool, aesthetics, usefulness, and perceived value. Parents from our Pediatric Parent Advisory Group (P-PAG) were asked to provide feedback on the length, stylistic elements, and highlight areas not addressed in the tools. The P-PAG was developed by ECHO and ARCHE and meets once a month to provide parental feedback on our tools and research activities. Research team meetings are held weekly to discuss the development of our tools. Our KT Tool Development Cycle can be viewed in **Figure 1**.

### Video

Five versions of the video were developed before the tool was ready for usability testing. The closed-captioned, English-language video is 5 minutes and 20 seconds in length and features a family of four (father, mother, older brother, and younger baby sister). The story begins with the brother, Sam, developing a cold which was later passed onto his younger sister, Kira. Unlike Sam who recovered quickly from the cold, Kira’s symptoms worsened, and she began exhibiting symptoms of bronchiolitis which included wheezing and skin sucking in around her neck and ribs. Following several attempts at managing Kira’s symptoms at home, the parents seek emergency care, resulting in a diagnosis of bronchiolitis. The video then outlines what bronchiolitis is, the symptoms associated with bronchiolitis, as well as the differences in how symptoms can manifest for the babies and older children. This is followed by general information on how bronchiolitis can be managed at home and when to seek professional help. Finally, the video describes what happens during an ED visit for bronchiolitis, if required.

The video was later entered into the Canadian Institutes of Health Research (CIHR) Institute of Human Development, Child and Youth Health (IHDCYH) Talks Video Competition (2019) and received special commendation.

### E-Book

In the 18-page eBook, a father narrates his experiences when his infant daughter, Rose, develops a cough, runny nose, and refuses to eat. Despite his best efforts to manage her symptoms at home, she does not get better. He calls the 24-hour health information line and is told to go to the ED. There, he learns that she has bronchiolitis. Throughout the story, the eBook describes common symptoms for infants and older children, risk factors, diagnosis methods, and common treatments. The story concludes with home care tips. To increase reader understanding, certain terms are defined in the appendix.

### Infographic

The bronchiolitis interactive infographic was developed under the same style as our suite of infographics. The style is unique to our research program and underwent dozens of iterations and two years of development. In order to ensure consistency across all pediatric conditions, the same style, design elements, animations, and characters are used.

The infographic looks similar to a webpage and allows users to scroll through the information, exploring at their own pace. The ability for parents to control what they view paced on their needs differentiates the interactive infographic from the video and e-Book. The information provided in the bronchiolitis infographic mirrors information that is provided in both the video and e-Book and is comprised of five major sections: (1) General Information about Bronchiolitis, (2) Here’s What You Can Do, (3) Treatment, (4) Useful Links, and (5) Contact Us. Within *General Information*, subheadings include what bronchiolitis is, common symptoms, and risk factors. Under *Here’s What You Can Do*, the tool provides information on what parents could do at home, when to seek emergency care, and what to do if they are confused. The infographic also includes quotes from parents who have had children affected by bronchiolitis.

### Surveys

Parents were recruited to participate in an electronic, usability survey (**Appendix E**) in two urban ED waiting rooms in the Edmonton area (Stollery Children’s Hospital and Northeast Community Health Centre). Surveys were informed by a systematic review of over 180 usability evaluations and comprised of 9, 5-point Likert items assessing: 1) usefulness, 2) aesthetics, 3) length, 4) relevance, and 5) future use. Parents were also asked to provide their positive and negative opinions of the tool via two free text boxes [20]. Members of the study team approached parents in the ED to determine interest and study eligibility. Study team members were available in the ED to provide technical assistance and answer questions as parents were completing the surveys.

### Data Analysis

Data were cleaned and analyzed using SPSS v.24. Descriptive statistics and measures of central tendency were generated for demographic questions. Likert answers were given a corresponding numerical score, with 5 being “Strongly Agree” and 1 being “Strongly Disagree” [21-22]. One-way analysis of variance (ANOVA) was used to determine whether there were statistically significant differences between the three tools. Open-ended survey data were analyzed thematically. A summary of the results was then shared with the creative team to inform the development of the final versions of the tools.

## Results

93 parents from 2 ED waiting rooms waiting rooms participated in this study. Of those, 33 reviewed the infographic, 30 reviewed the eBook, and 30 reviewed the video.

In general, feedback for the bronchiolitis video and interactive infographic were positive, with participants giving the tools scores between 4.00 (*agree*) and 5.00 (*strongly agree*). Mean responses for the eBook varied more and ranged from 3 (*neither agree nor disagree*) to 5 (*strongly agree*). When asked about the usefulness of the tools, all parents strongly agreed or agreed that the tools were useful. High scores were also reported for relevance, simplicity, and aesthetics. Parents also indicated all tools could be used without written instructions or additional help.

A significant difference is seen in the means regarding appropriateness of length; with a mean of 4.09 for the infographic length, 3.33 for the e-Book, and 4.30 for the video. The video and infographic also scored higher than the e-Book when parents were asked about plans for future use and decision-making, although differences were not statistically significant. Nonetheless, parents strongly agreed and agreed that they would recommend the tools to their friends. Table 2 displays the mean responses to the 9 usability questions based on tool.

**Table 1.** Demographic characteristics of parents who assessed the usability of the bronchiolitis e-tools (N=93)

| Characteristic | n (%) |
| --- | --- |
| Sex |  |
| Female | 69 (74.2) |
| Male | 24 (25.8) |
| Age |  |
| Less than 20 | 1 (1.1) |
| 20-29 years | 20 (21.5) |
| 30-39 years | 40 (43.0) |
| 40-49 years | 18 (19.4) |
| 50-59 years | 10 (10.8) |
| 60 years and older | 4 (4.3) |
| Marital Status |  |
| Married/Partnered | 71 (76.3) |
| Single/Separated/Divorced/Widowed | 22 (23.7) |
| Education |  |
| Some high school | 10 (10.8) |
| High school diploma | 19 (20.4) |
| Some post-secondary | 16 (17.2) |
| Post-secondary certificate/diploma | 20 (21.5) |
| Post-secondary degree | 15 (16.1) |
| Graduate degree | 10 (10.8) |
| Other | 2 (2.2) |
| Household Income |  |
| Less than \$25,000 | 8 (8.6) |
| \$25,000-\$49,000 | 26 (28.0) |
| \$50,000-\$74,000 | 15 (16.1) |
| \$75,000-\$99,000 | 17 (18.3) |
| \$100,000-\$149,000 | 13 (14.0) |
| \$150,000 and over | 9 (9.7) |
| Prefer not to answer | 3 (3.2) |
| Number of Children in the Family |  |
| 1 | 23 (24.7) |
| 2 | 37 (39.8) |
| 3 | 17 (18.3) |
| 4 | 11 (11.8) |
| 5+ | 4 (5.4) |

**Table 2.** Means (SD) of participant responses to the usability survey *p < 0.05.

| **Usability Measures** | **Video** | **Infographic** | **e-Book** |
| --- | --- | --- | --- |
| It is useful. | 4.53 (0.82) | 4.39 (0.50) | 4.37 (0.67) |
| It provides information that is relevant to me as a parent. | 4.40 (0.56) | 4.33 (0.60) | 4.30 (0.79) |
| It is simple to use. | 4.60 (0.50) | 4.27 (0.72) | 4.23 (0.86) |
| I can use it without written instructions or additional help. | 4.50 (0.57) | 4.21 (0.78) | 4.30 (0.84) |
| Its length is appropriate. | 4.30 (0.53) | 4.09 (0.95) | 3.33 (0.96) |
| It is aesthetically pleasing (i.e. images, colours, etc.). | 4.53 (0.57) | 4.42 (0.67) | 4.37 (0.61) |
| It helps me make decisions about my child's health. | 4.23 (0.63) | 4.22 (0.61) | 3.97 (0.72)* |
| I would use it in the future. | 4.40 (0.67) | 4.24 (0.79) | 3.97 (0.69) |
| I would recommend it to a friend. | 4.43 (0.57) | 4.38 (0.61) | 4.10 (0.66) |

\*p < 0.05.

In the open text boxes, parents described the bronchiolitis video as “clear”, “concise”, and “visually appealing”. One parent stated that they liked that the information was presented in a manner that was not too complicated. Another parent said, “This is an awesome tool that I would use. It should be posted in waiting rooms and on tv commercials”.

Parents described the infographic as being very “well done” and that they appreciated that there was work and research being done in the pediatric knowledge translation space. Finally, parents described the e-Book as being a good resource to go through with their children and would be a valuable method of teaching them about illnesses. They liked that the book provided details about what to expect in the emergency department and were appreciative of the summary lists included in the book. Furthermore, parents described the storyline of the book as a way to teach children about empathy as well.

## Conclusions

We developed and evaluated the usability of three innovative, arts-based, digital tools for parents with children with bronchiolitis. Using comprehensive methods that include (1) synthesis of evidence from the literature; (2) collection of our own evidence through interviews with parents who have had children with bronchiolitis; (3) content outline development; (4) tool design and development; (5) several rounds of revisions and modifications with stakeholders and the study team, and (6) usability testing with our end-users, we developed several tools that were highly rated and met the information needs of parents. High scores across all usability items for all tools suggest that using multi-method development processes can result in tools that are useful, relevant, understandable, and will be used by the intended audience.

Our tools are easily accessible and can be viewed across a broad range of devices, including desktop computers, mobile phones, and tablet devices. Providing a means for quick and convenient access ensures that parents can obtain information about how to manage their child’s condition quickly and effective. Such resources may reassure parents, support them in caring for their children at home, and help them make evidence-based decisions about accessing health care services. This may also alleviate the burden on the health system.

**The tools can be found here: http://www.echokt.ca/tools/bronchiolitis/**

Note: Our KT tools are assessed for alignment with current, best-available evidence every two years. If recommendations have changed, appropriate modifications are made to our tools to ensure that they are up-to-date [23].

## Data Availability

Data referred to in the manuscript are not available for access. Data are stored at on secured servers at the University of Alberta to protect participant privacy and confidentiality.

## Author Contributions

This study was conducted under the supervision of Drs. Shannon D. Scott (SDS) and Lisa Hartling (LH), PIs for **translation Evidence in Child Health to enhance Outcomes** (ECHO) Research and the **Alberta Research Centre for Health Evidence**, respectively. Both PIs designed the research study and obtained research funding through Translating Emergency Knowledge for Kids (TREKK) Networks of Centres of Excellence of Canada (NCE), Stollery Children’s Hospital Foundation, and the Women and Children’s Health Research Institute.

SS developed and supervised all aspects of tool development and evaluation.

LH contributed to all aspects of tool development and evaluation and supervised the systematic review of parent experiences and information needs.

Alyson Campbell and Samantha Louie-Poon conducted and analyzed qualitative interviews with parents.

Tony An developed the infographic.

Tabatha Plesuk and Hyelin Sung collected usability data.

Anne Le analyzed usability data.

All authors contributed to the writing of this technical report and provided substantial feedback.

This work was funded by:

**Networks of Centres of Excellence:**

- Klassen, T., Hartling, L., Jabbour, M., Johnson, D., & Scott, S.D. (2015). Translating emergency knowledge for kids (TREKK). Networks of Centres of Excellence of Canada Knowledge Mobilization Renewal ($1,200,000). January 2016 – December 2019.

**Women’s and Children Health Research Institute**

- Scott, S.D & Hartling L. (2016). Translating Emergency Knowledge for Kids renewal. Women and Children’s Health Research Institute (matched dollars, $150,000). 2016/04- 2019/04.

## Other Outputs from this Project

## Appendices

## Appendix A Interview Guide

Parents will be interviewed to understand their experience having a child with bronchiolitis. Semi-structured interviews will be conducted with parents in order to get their “narrative” or experiences. The following questions will be used to guide these interviews. Being true to semi-structured interview techniques, interview questions will start broad and then move to the more specific.

1. Tell me about your experience having your child experience bronchiolitis.
2. Tell me about your child that was ill. How old is your child? How was your child ill? Has your child previously bronchiolitis?
3. How did you feel during this experience?
4. Tell me how prepared you felt during the experience. Did you have all of the information you needed to make decisions about when to seek healthcare? Tell me more about that.
5. What did you do to manage symptoms of bronchiolitis? (any techniques you used, for example, giving Tylenol, talking with family/friends, etc.)
6. How was your experience in the ED? Tell me about it.
7. Tell me about your child being diagnosed with bronchiolitis – were any tests done? Any medications ordered?
8. What strategies were put in place by health care professionals to help your child? (for example, giving/prescribing medication). Did they ask you to do anything?
9. How did your child manage the experience? How did you feel about the outcome of this situation?
10. If presented with the same situation again, would you do anything differently? If so, please tell me.

## Appendix B Video Images

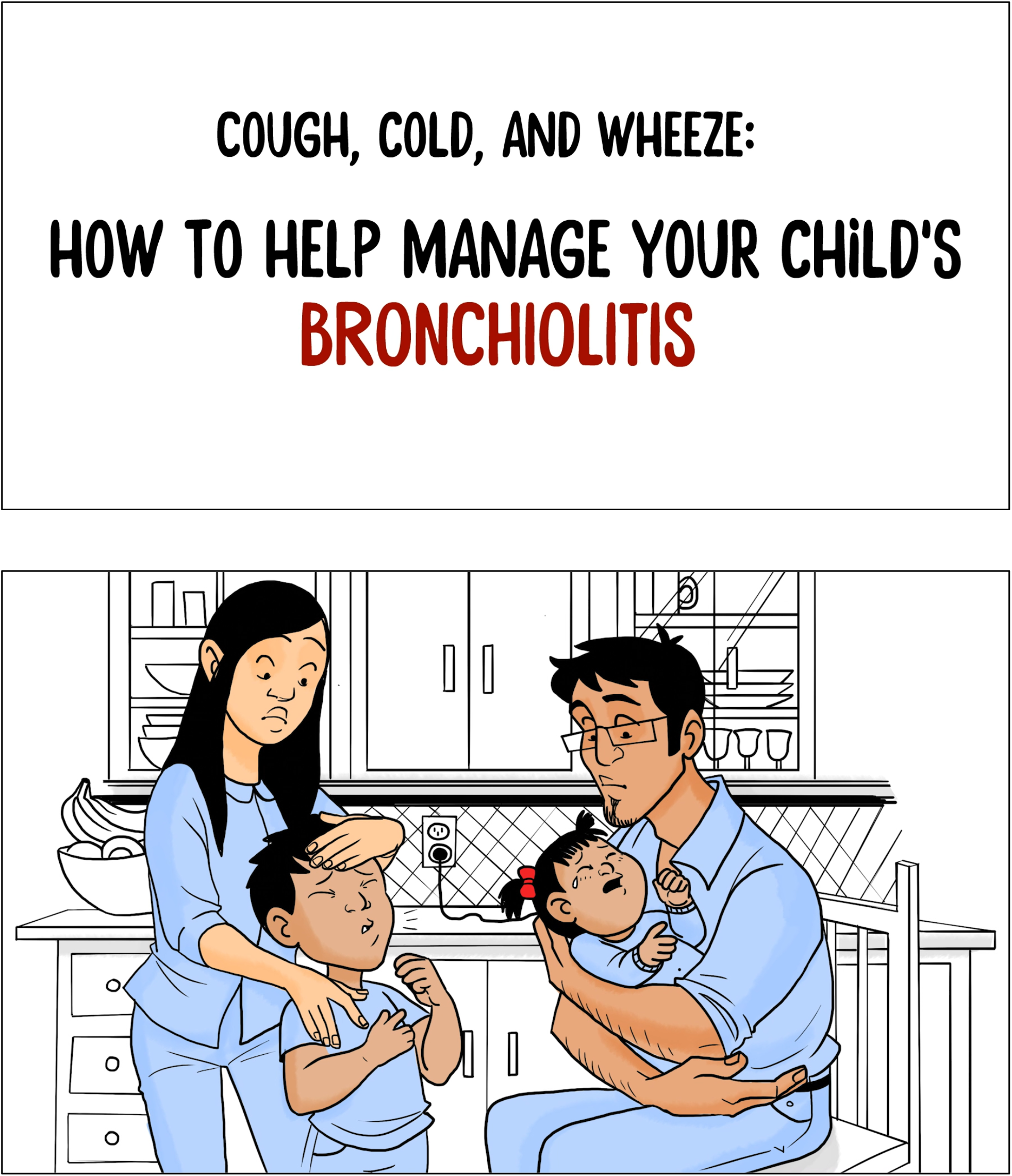

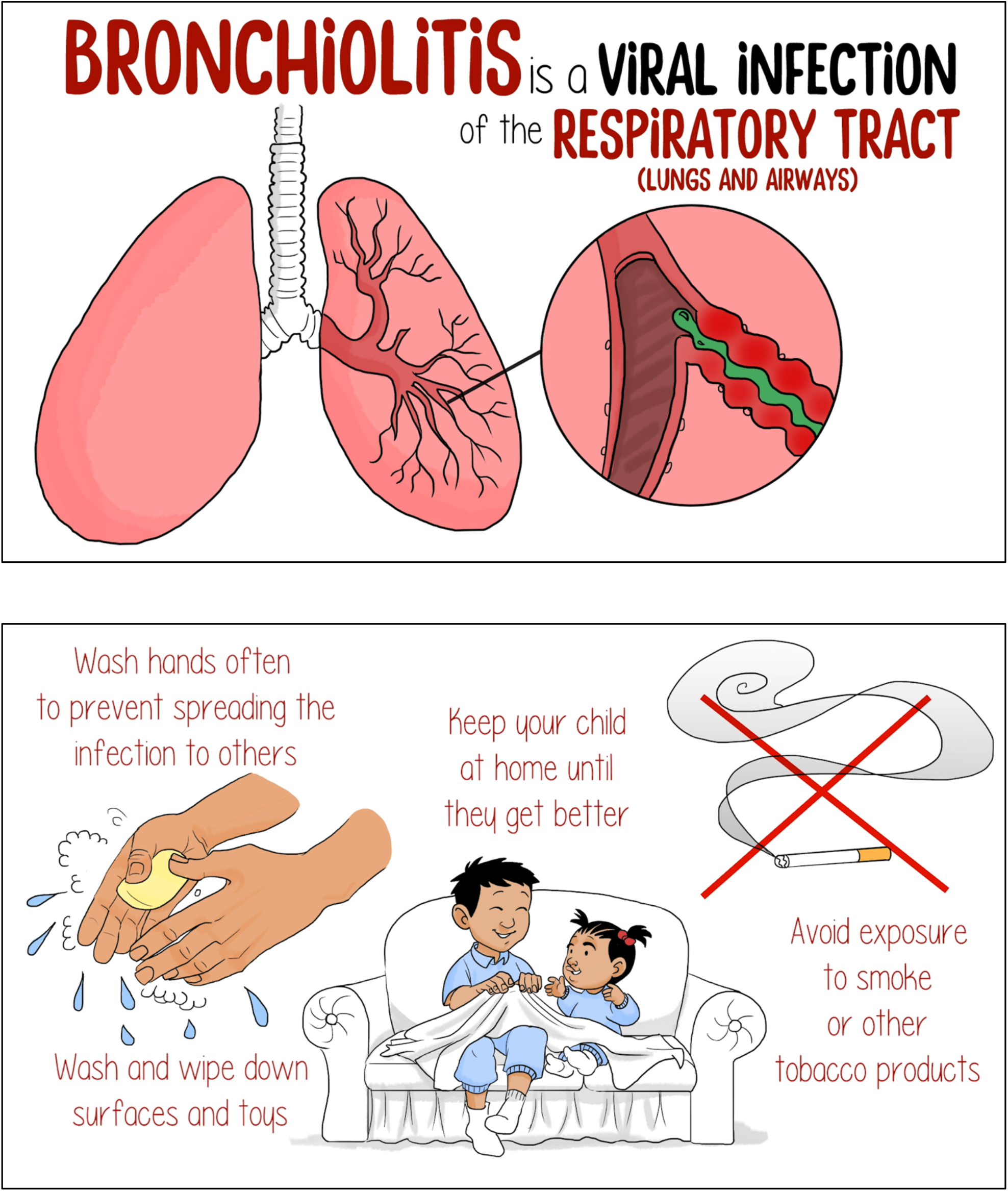

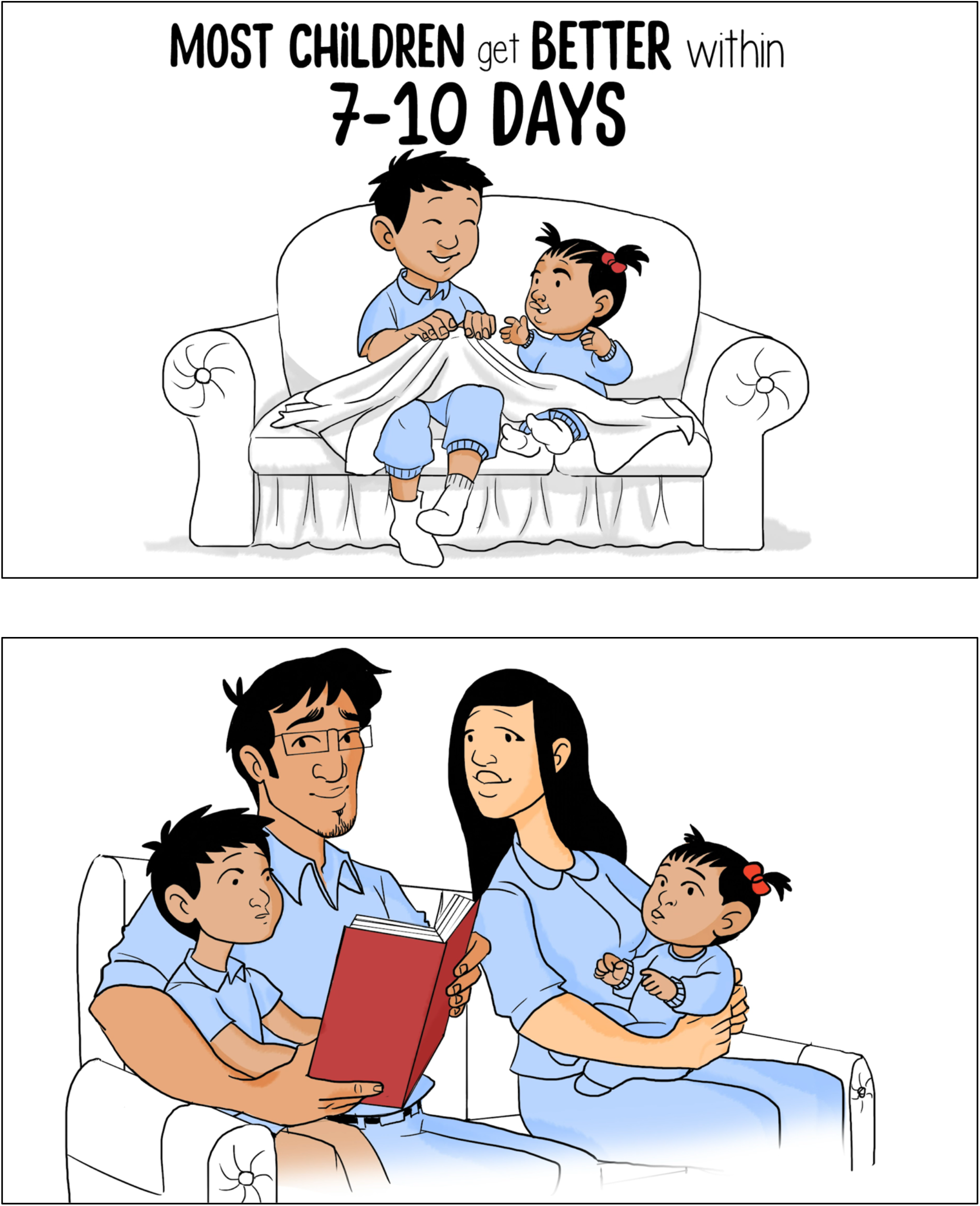

## Appendix C e-Book Images

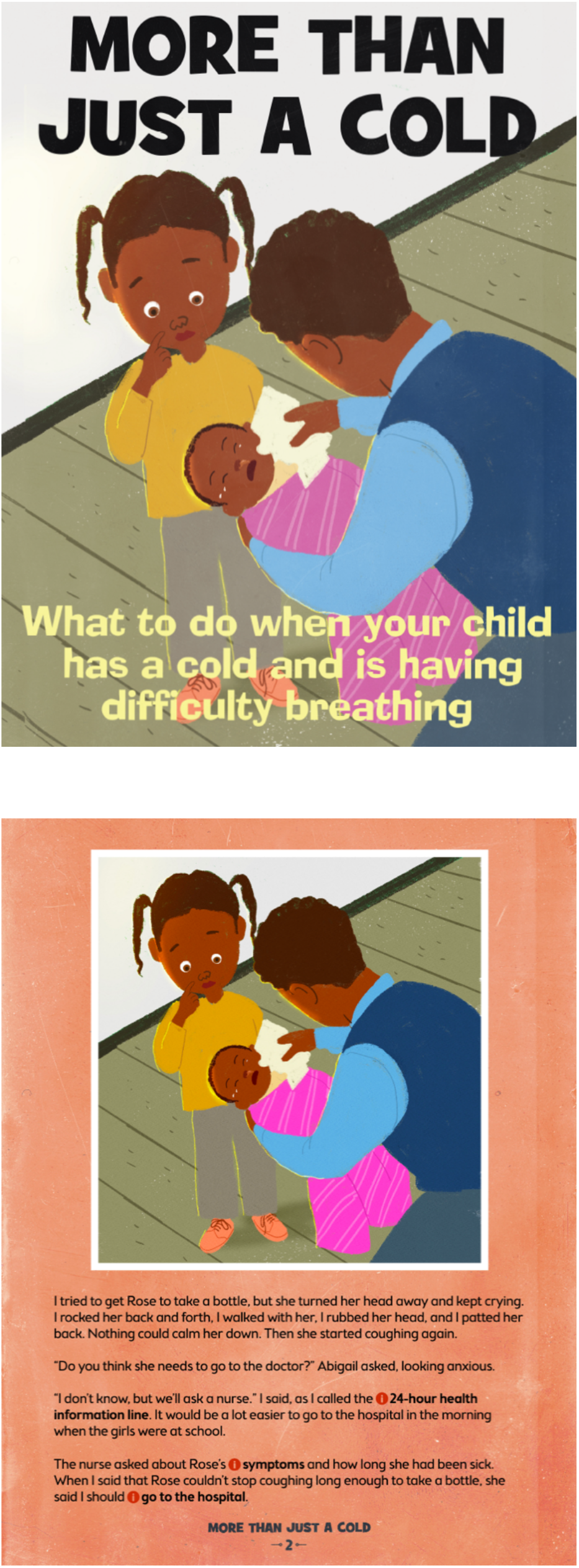

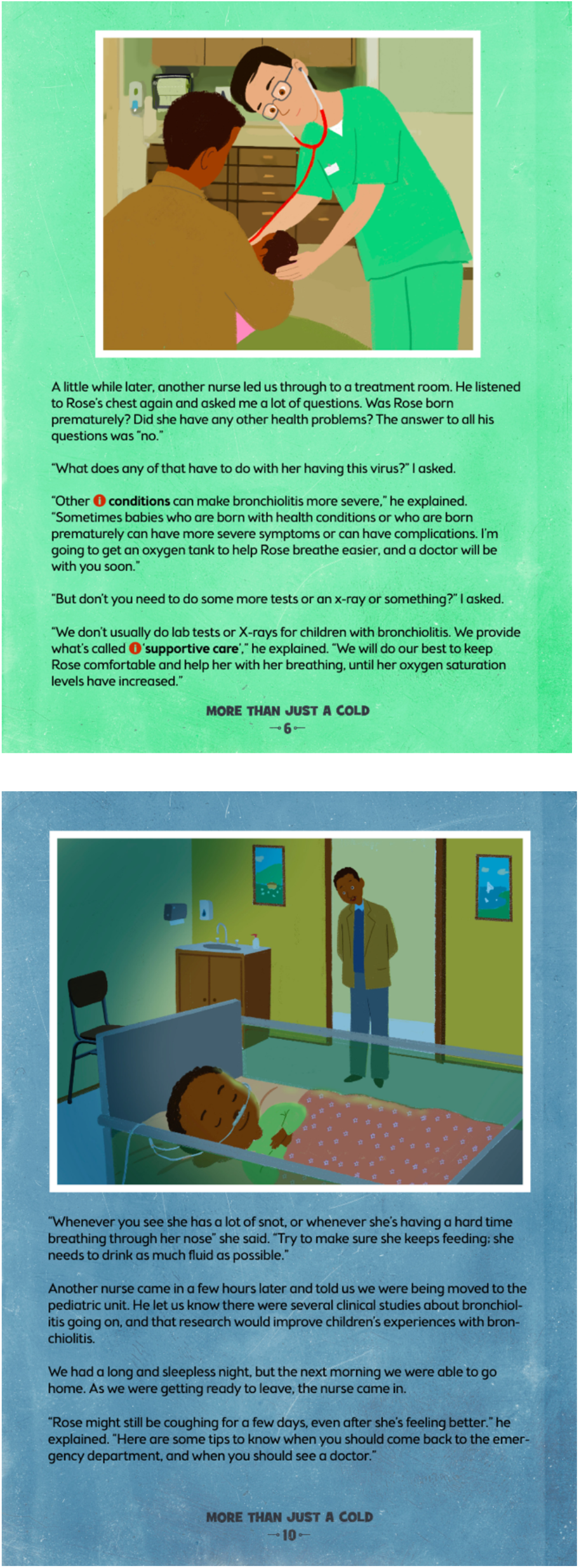

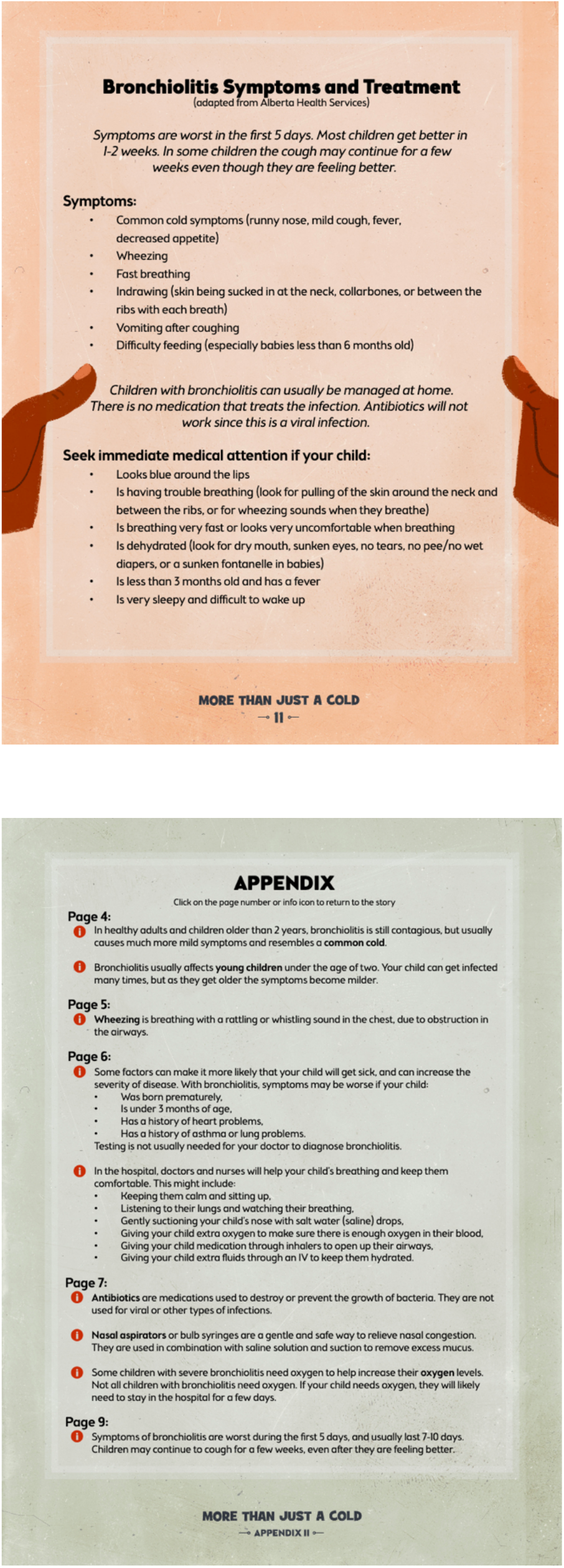

## Appendix D Infographic Images

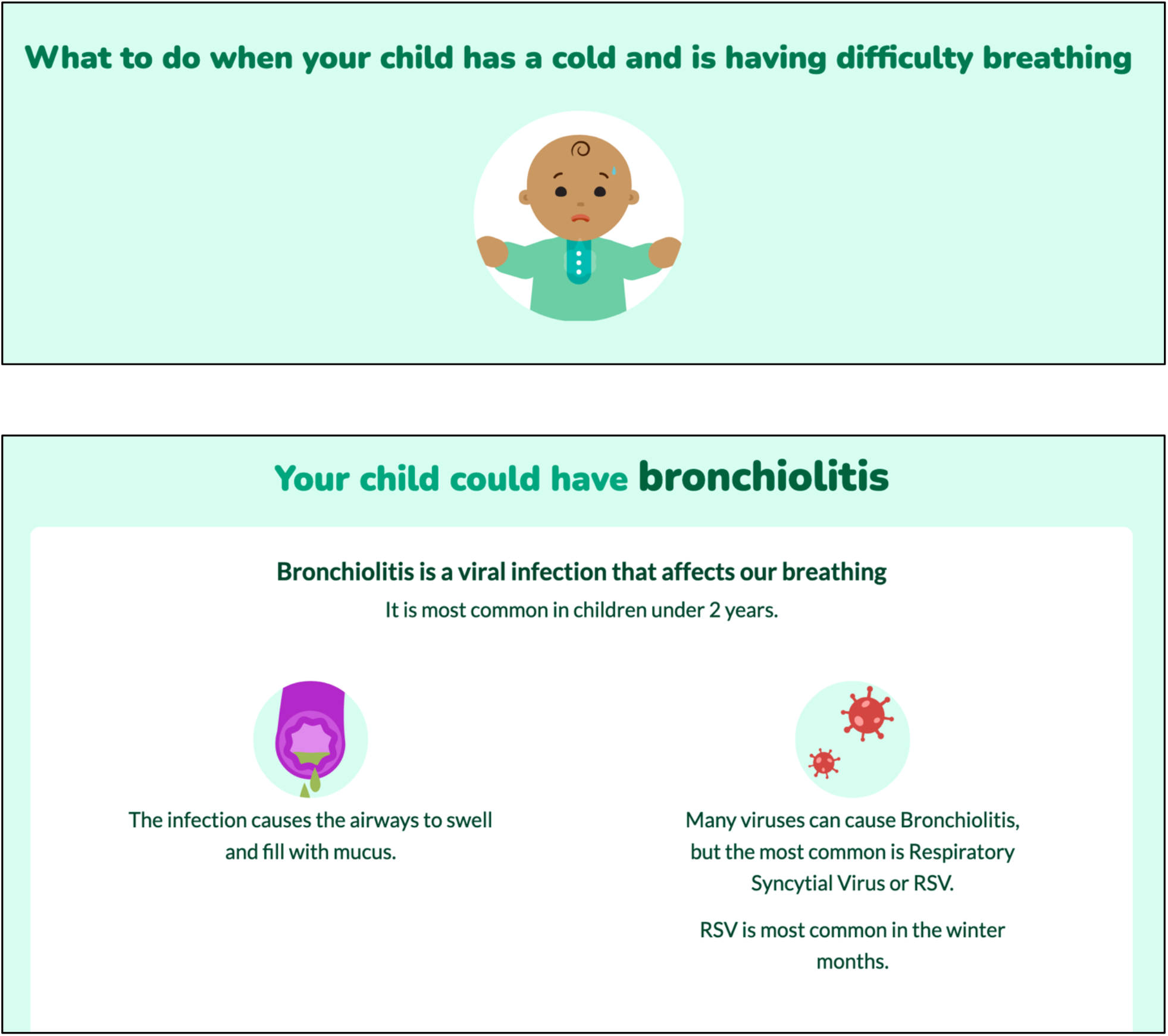

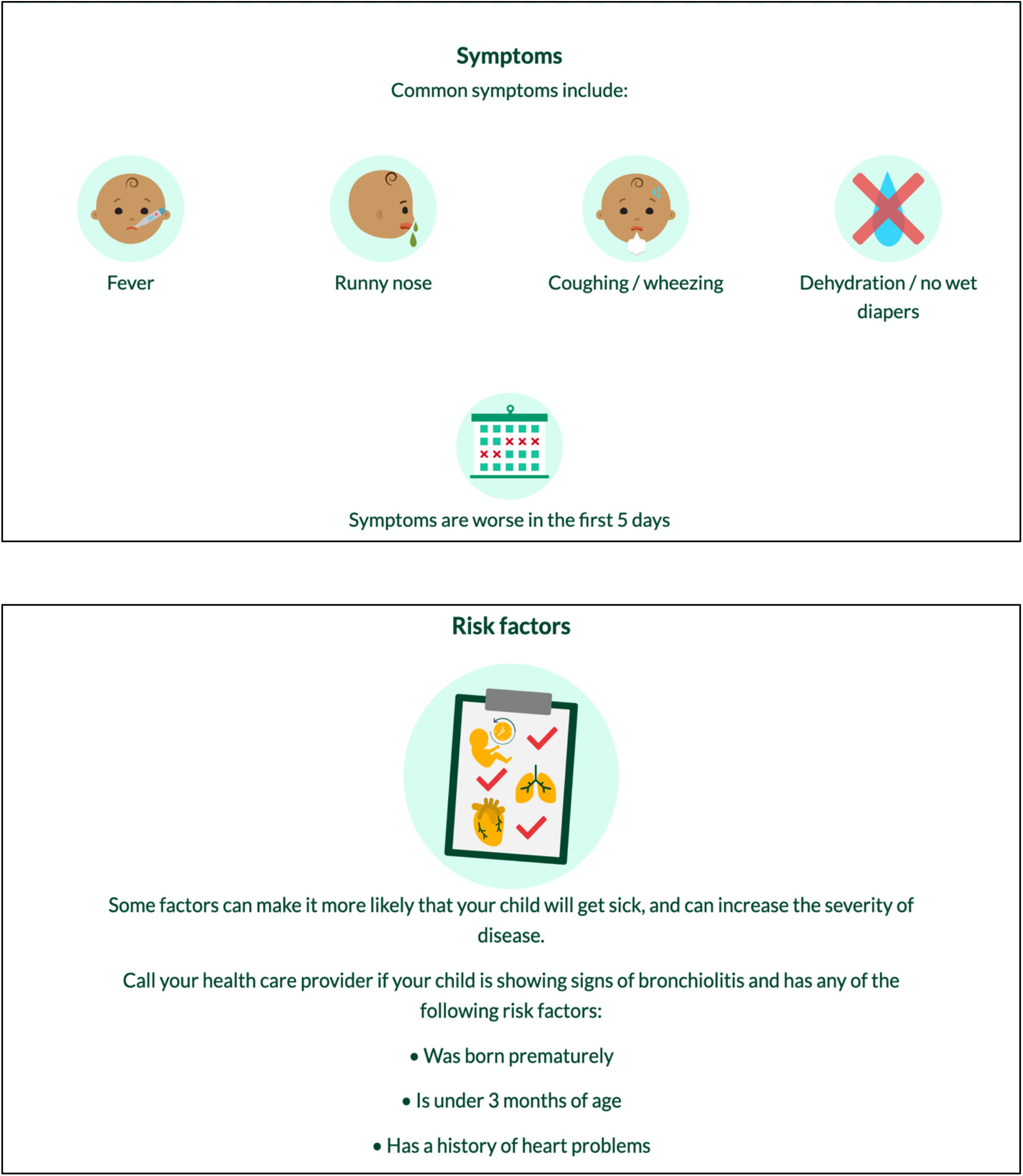

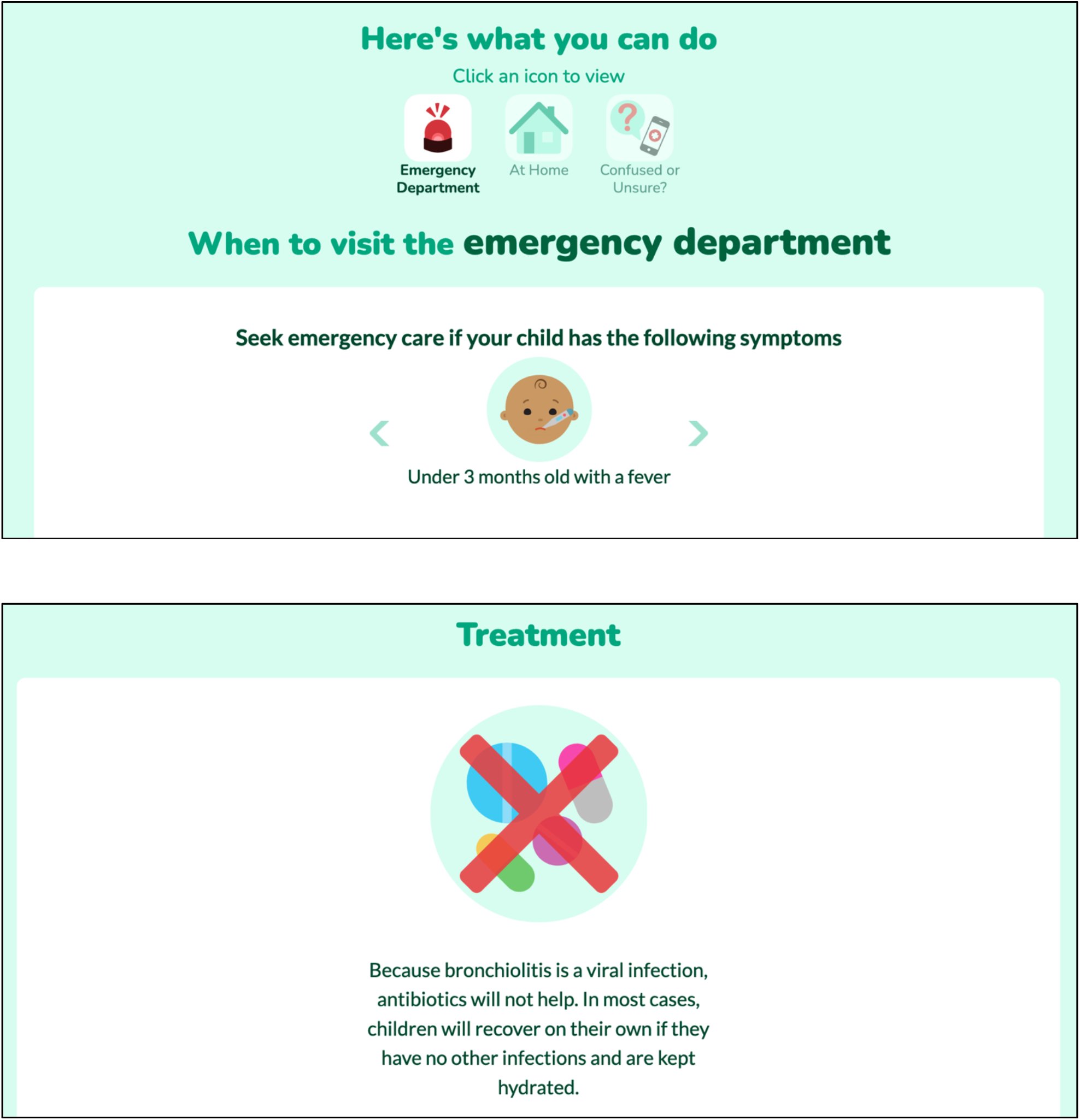

## Appendix E Usability Survey

SECTION 1: Demographics

1) What is your gender?
  □ Male
  □ Female
3) What is your Age?
  □ Less than 20 years old
  □ 20-30 years
  □ 31-40 years
  □ 41-50 years
  □ 51 years and older
7) What is your Marital Status?
  □ Married
  □ Single
5) What is your gross annual household income?
  □ Less than $25,000
  □ $25,000-$49,999
  □ $50,000-$74,999
  □ $75,000-$99,999
  □ $100,000-$149,999
  □ $150,000 and over
6) What is your highest level of education?
  □ Some high school
  □ High school diploma
  □ Some post-secondary
  □ Post-secondary certificate/diploma
  □ Post-secondary degree
  □ Graduate degree
  □ Other
7) How many children do you have?
8) How old are your children?

SECTION 2: Assessment of attributes of the arts-based, digital tools

***\*participant is randomized to view 1 of 3 digital tools then automatically directed to the survey***

1. It is useful. [5-point Likert Scale]
2. It provides information that is relevant to me as a parent. [5-point Likert Scale]
3. It is simple to use. [5-point Likert Scale]
4. I can use it without written instructions or additional help. [5-point Likert Scale]
5. Its length is appropriate. [5-point Likert Scale]
6. It is aesthetically pleasing (i.e., images, colours, etc.). [5-point Likert Scale]
7. It helps me to make decisions about my child’s health. [5-point Likert Scale]
8. I would use it in the future. [5-point Likert Scale]
9. I would recommend it to a friend. [5-point Likert Scale]
10. List the most negative aspects: [open text]
11. List the most positive aspects: [open text]

## Appendix F Project Timeline

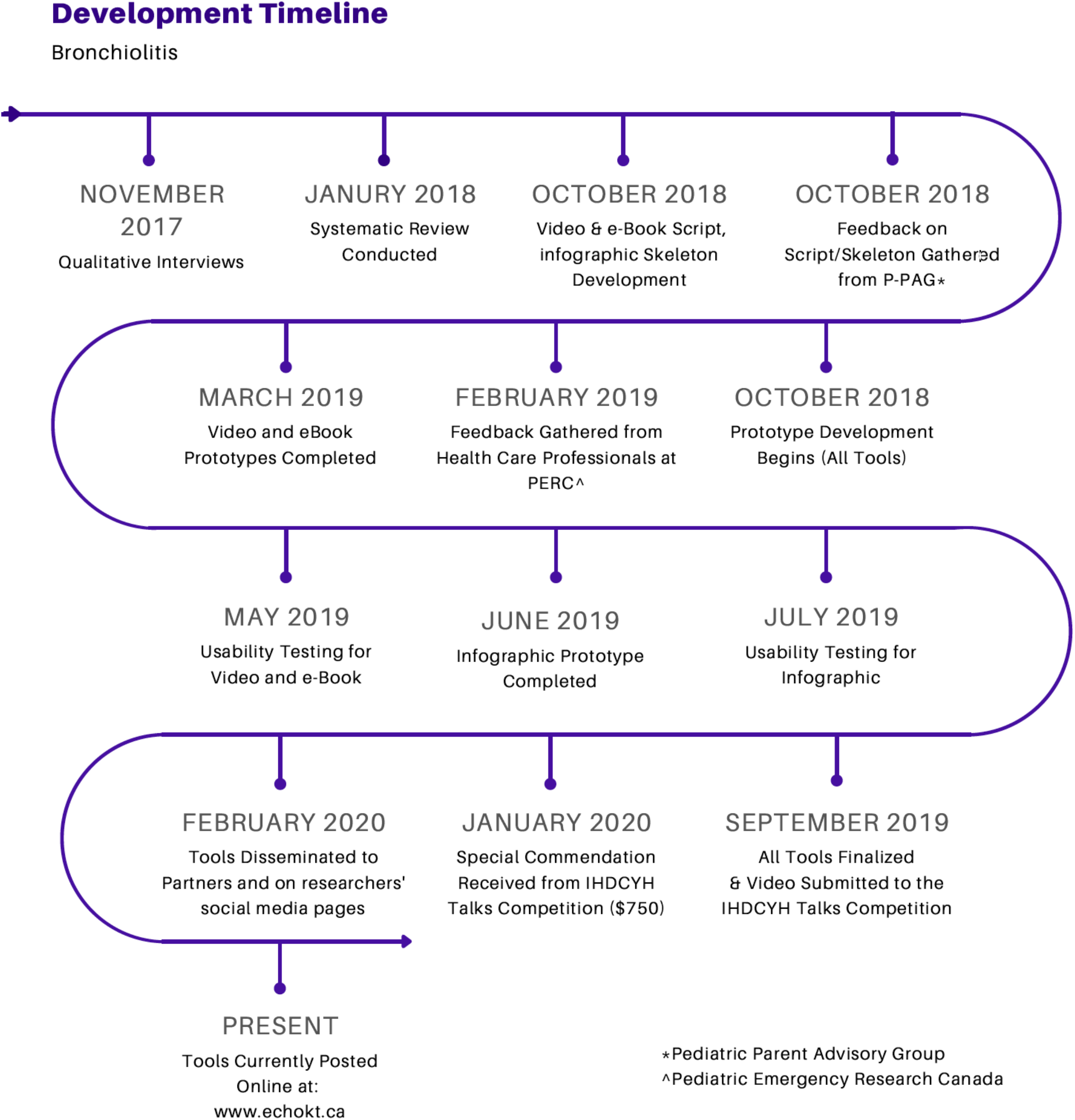

